# Computation of Expected Epidemic Duration

**DOI:** 10.1101/2023.04.24.23289026

**Authors:** Alejandro Alarcón González, Guillermo A. Pérez

## Abstract

This paper discusses the mean duration of a closed epidemic modeled by a discrete-time Markov chain. We develop a methodology for the efficient computation of the quantity of interest. The Markov chain model in consideration is bivariate, and is formally handled. We derive explicit terms for the probability to transition from one state to another, and prove that the chain is absorbing. The computation of the mean duration is translated to the computation of the expected hitting times to the set of absorbing states. We use the theory of absorbing Markov chains to derive a matrix formulation that gives way to an efficient algorithm to solve for the expected hitting times. This approach is instantiated in the form of a concrete algorithm, which is further optimized by using dynamic programming. Finally, we have implemented the method and tested it against the use of simulations to estimate mean durations.

## 1. Introduction

Stochastic epidemic models aim to reflect the nature of the contact and transmission processes involved in the spread of disease in a population. However, their analysis tends to be difficult, which motivates the need for mathematically tractable yet realistic models. This is the context where Markovian epidemic models find their relevance. Markov models are stochastic systems for which the future depends only on the current state of the system. They rely on a matured mathematical theory (see for instance [1] and [2]) and enjoy a good deal of popularity in the biological sciences (see for instance [3] and [4]).

A substantial share of effort in mathematical epidemiology has been devoted to the computation of quantities of interest, with (perhaps) the most famous example being the basic reproduction number *R*_0_ expressed as the leading eigenvalue of the next generation matrix [5]. The quantity we consider in this work is the duration of the stochastic epidemic. That is, the time that elapses till the extinction of the disease. For the class of Markovian epidemic models, Bailey [6] analyzed the Reed-Frost and Greenwood models and derived probability generating functions for the duration of the epidemic in those models. Keeling and Ross [7] in turn addressed the computation of the expected duration in a continuous-time Markovian model.

In this paper, we analyze a discrete-time Markovian epidemic model in a closed population. It is made by discretizing time in a simple deterministic epidemic model and then turning it stochastic with tools from basic probability theory. This derivation follows the work done by Abrams et al. [8], although we take as a basis the much simpler deterministic SIR model [9]. The resulting Markovian epidemic model resembles the chain-binomial constructions of Reed-Frost and Greenwood [6], but with more realistic features: e.g. the time during which an individual remains infectious is not just an instant. However, it is simple enough to allow for a clear exposition of the mathematical and algorithmic treatment herein developed.

Similarly to Black and Ross [10], we analyze the transition matrix of a Markovian epidemic model and then state our research problem in terms of a matrix formulation. Similarly as well, the method we devolop yields the exact value for the quantity of interest. However, the class of models they discuss stem from an event-driven perspective. That is, the only two valid transitions making up such models correspond to an infection and a recovery event. Such event-driven approach is frequently used (see for instance [11, 7, 9]). Instead, the model we analyze stems from a probabilistic interpretation of an Euler-forward discretization of a system of differential equations, which makes it discrete-time and with a constant time step size. Resulting from this construction the number of infectious and recovery events per time step can be very large, as it will be explained in the fourth section.

We address the problem of computing the mean duration of the epidemic in the model we consider. Interestingly, our algorithm gives us these values from all possible starting states. This problem translates to the well known calculation of the mean hitting times in a discrete-time Markov chain [1], which is a system of linear equations. We emphasize that our method can be used to analyze models with more compartments and with contact structure.

### Contributions

As a first contribution, we derive an explicit formula for the probability to transition from one state to another in the Markovian model we consider. Then, based on the latter we derive a matrix formulation of the mean hitting times. By analyzing the structure of this matrix we are able to give an expression for the hitting times that already yields an efficient algorithm. Our method is further optimized by using dynamic programming, a mathematical optimization technique that resembles the use of a Pascal triangle to compute binomial coefficients.

Finally, we have implemented the method and tested it against the use of simulations to estimate mean durations.

## 2. Preliminaries

In this section we give a definition of Markov chains and recall some of their basic properties. We mostly follow the notation from [1] for stochastic processes and Markov chains.

A (finite-state discrete-time) *stochastic process* is a sequence of random variables *X*_*t*_, for *t* ∈ ℕ, all defined on a common probability space and taking values from the same finite set *Q* of states. We write (*X*_*t*_)_*t*≥0_ for the sequence of variables. Additionally, we often make implicit use of the probability space (Ω, ℱ, Pr) induced by the stochastic process. In particular Ω, the set of outcomes, consists of all sequences of possible values for the random variables. Then, ℱ is a *σ*-algebra over Ω and Pr is a probability measure on ℱ. In this probability space, a *random variable X* is a function *X* : Ω × ℕ → *Q*. Usually, we write Pr(*X*_*t*_ = *i*) to denote Pr(*{ω* ∈ Ω : *X*(*ω, t*) = *i}*).

### 2.1. Markov chains

A (finite-state discrete-time) *Markov chain* is a stochastic process (*X*_*t*_)_*t*≥0_ such that the sequence starts with an initial distribution over *Q*, specified by ***λ*** = (*λ*_*i*_ : *i* ∈ *Q*), and satisfies the *Markov property*. That is, for all *t* ∈ ℕ and all *i*_0_, …, *i*_*t*+1_ ∈ *Q* we have that the probability of *X*_*t*+1_ taking the value *i*_*t*+1_ depends only on the value of *X*_*t*_. In symbols:

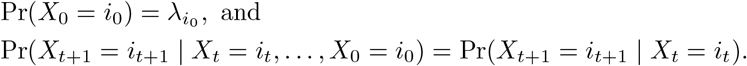

Note that this definition guarantees the existence of the so-called *transition matrix* ***P*** = (*p*_*ij*_ | *i, j* ∈ *Q*), where:

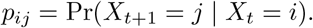

The matrix ***P*** is *stochastic*, i.e., 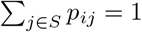, for all *i* ∈ *Q*.

We shall make use of the following characterization of the Markov property based on the transition matrix (see, e.g., [1, Theorem 1.1.1]).

#### Theorem 1 (Markov property)

*Let* (*X*_*t*_)_*t*≥0_ *be a stochastic process. It is a Markov chain if and only if for all n* ∈ ℕ *and all i*_0_, …, *i*_*n*_ ∈ *Q the following holds*.

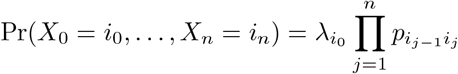

Later, it will be useful to consider several probability measures depending on fixed initial states for the Markov chain. For this purpose, let Pr_*i*_ denote the probability measure obtained by setting ***λ*** so that *λ*_*i*_ = 1 and *λ*_*j*_ = 0 for *j* ≠ *i*. Similarly, let E_*i*_[·] for the expectation operator with respect to Pr_*i*_.

#### 2.1.1. Absorbing Markov chains

We say that state *j* is *reachable* from state *i* if and only if for some *t* ∈ ℕ:

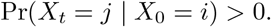

A state *i* ∈ *Q* is *absorbing* if and only if *p*_*ii*_ = 1. If the set of absorbing states *A* of a Markov chain (*X*_*t*_)_*t*≥0_ is not empty and *A* is reachable from all states, we say that (*X*_*t*_)_*t*≥0_ is an *absorbing Markov chain*. In an absorbing Markov chain, a state that is not absorbing is *transient*.

The transition matrix of an absorbing Markov chain has special properties. Consider an ordering of *Q* such that the *t* transient states are first, followed by the *r* absorbing states. Now, the transition matrix will have the following *canonical form*.

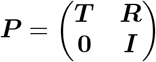

Above, ***I*** is a *r* × *r* identity matrix; **0**, a *r* × *t* matrix of zeros; ***R***, a nonzero *t* × *r* matrix; and ***T***, a *t* × *t* matrix.

##### Theorem 2.

*Consider an absorbing Markov chain* (*X*_*t*_)_*t*≥0_. *The matrix* ***I*** – ***T*** *has an inverse*.

The result above is well known. It is, for example, stated and proven in [2, Theorem 11.4]. The inverse of ***I*** − ***T*** is commonly written ***N*** and called the *fundamental matrix*.

#### 2.1.2. Expected hitting times

Let *A* ⊆ *Q* be a set of *target states*. We write *τ*_*A*_ to denote the first *hitting time* of a state in *A*. More formally, *τ*_*A*_ is the random variable:

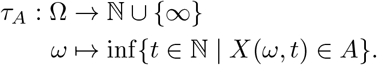

Note that *τ*_*A*_ can take countably many values only and they all are nonnegative. Hence, its expectation, denoted 𝔼[*τ*_*A*_], satisfies the following (see also [1, Section 1.3]).

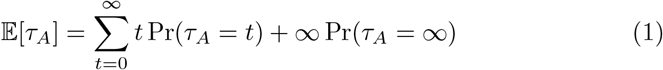

Let *i* ∈ *Q* be a state and write 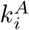 for the value E_*i*_[*τ*_*A*_]. The following characterization of the expected hitting times will be useful later. The result is well known and can be found, for instance, in [1, Theorem 1.3.2].

##### Theorem 3.

*The vector of expected hitting times* 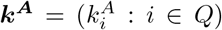 *is the minimal (w*.*r*.*t. the product order) nonnegative solution to the following system*.

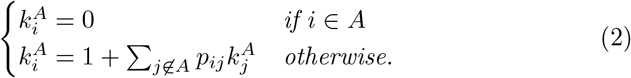

### 2.2 Expected hitting times in absorbing Markov chains

For absorbing Markov chains, the probability that the process reaches an absorbing state is one. In fact, the probability of not having reached it decreases exponentially (see, e.g., [2, Proof of Theorem 11.3]).

#### Lemma 1.

*Consider an absorbing Markov chain* (*X*_*t*_)_*t*≥0_ *with absorbing set of states A* ⊆ *Q. There exist r* ∈ (0, 1) *and m* ∈ ℕ *such that for all states i* ∈ *Q and all n* ∈ ℕ *the following holds*.

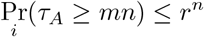

Proof: The argument hinges on the facts that the Markov chain has a finite state space and that it is absorbing. Indeed, from each state *i* there is some absorbing state *j* that is reachable. Let *m*_*i*_ be the minimum number of steps needed for *i* to reach *j*. That is, *m*_*i*_ is the minimal *t* ∈ ℕ such that Pr_*i*_ (*X*_*t*_ ∈ *A*) *>* 0. Now, define *m* = min_*i*∈*Q*_ *m*_*i*_ and *r* = 1 − min_*i*∈*Q*_ *r*_*i*_ where 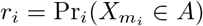. Observe that 0 *< r <* 1. Now, using the Markov property, we get that for all *i* ∈ *Q* the probability of not reaching an absorbing states after 2*m* steps is at most *r*^2^. In symbols: for all *i* ∈ *Q* we have Pr_*i*_ (*τ*_*A*_ ≥ 2*m*) ≤ *r*^2^. The claim then follows by induction.

Using the lemma above, we can prove that in absorbing Markov chains the expected hitting times are always finite.

#### Theorem 4

*Consider an absorbing Markov chain* (*X*_*t*_)_*t*≥0_ *with absorbing set of states A* ⊆ *Q. For all i* ∈ *Q we have that* Pr_*i*_(*τ*_*A*_ = ∞) = 0 *and* 𝔼_*i*_[*τ*_*A*_] *<* ∞.

Proof: For the first part of the claim, we define, for each *n* ∈ ℕ, the event *A*_*n*_ = *{ω* ∈ Ω : *τ*_*A*_(*ω*) ≥ *mn}*, where *m* is as in Lemma 1. Observe that the events are monotone decreasing, i.e. *A*_ℓ_ *⊇ A*_*n*_ for all ℓ *< n*, and that lim_*n*→∞_ *A*_*n*_ = *{ω* ∈ Ω : *τ*_*A*_(*ω*) = ∞*}*. Using *σ*-additivity and Lemma 1 we get, for all *i* ∈ *Q*, the following.

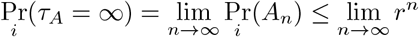

Since the last limit converges to 0, we are done.

For the second part of the claim, because of the first part together with Equation (1), it suffices to argue that 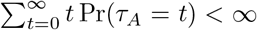. To make use of Lemma 1, we split the series into a finite sum of subseries, one per residue class modulo *m*. Formally, we have that the following hold.

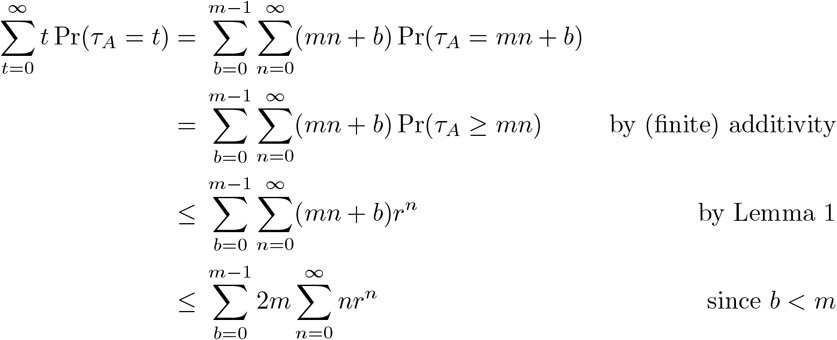

To conclude, we can use d’Alambert’s test (a.k.a. the limit test) to confirm that the series 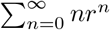 converges.

As a final result regarding the expected hitting time of absorbing Markov chains we observe that the above theorem can be obtained via linear algebra. In fact, a formula for the vector of expected hitting times exists in terms of the fundamental matrix (see, e.g., [2, Theorem 11.5]) and a vector **1** of all ones.

#### Theorem 5.

*Consider an absorbing Markov chain* (*X*_*t*_)_*t*≥0_ *with absorbing set of states A* ⊆ *Q and fundamental matrix* ***N*** *and let* 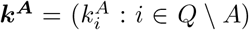 *be the vector of expected hitting times. Then*, ***k***^***A***^ = ***N* 1**.

In our developments below, we shall refine the above result for our specific transition and fundamental matrices.

## 3. SIR Models: From ODEs to Probabilities

Deterministic SIR Models are simple mathematical models of the spreading of infectious diseases. In them, a *population* of size *N* ∈ ℕ is partitioned into *compartments* with labels: *S* for susceptible, *I* for infectious, and *R* for recovered. As illustrated in Figure 1, people may move between compartments following time-dependent dynamics which are usually prescribed by ordinary differential equations (ODEs). Hence, we write *S*(*t*), *I*(*t*), and *R*(*t*) to highlight the fact that these values are function of time *t* ≥ 0. Below, we first state the deterministic SIR model for the case of a closed population [9], i.e., births, deaths or infections resulting from contacts with individuals from outside the population are not being considered. (The values *β* and *γ* are explained in the sequel.) Then, we derive a stochastic discrete-time version thereof based on Bailey’s chain binomial [6].

**Figure 1:**
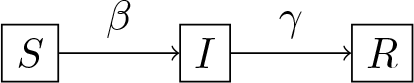
Overview of the flow of individuals in the SIR model: Following a disease infection, susceptible individuals (S) move to an infectious state (I) in which they can infect others. Such infectious individuals will recover over time.

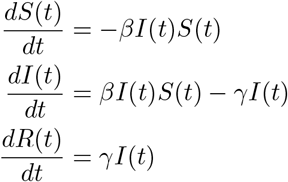

### 3.1. Towards a stochastic SIR model

When an infectious individual makes contact with a susceptible individual, there is some probability that such contact will lead to disease transmission. This probability times the contact rate is denoted by *β*^*\**^, and we take it to be irrespective of the specific susceptible-infectious pair. Furthermore, we define the *force of infection*, denoted by Λ(*t*), as the rate for a susceptible individual to become infected at time t. Assuming *homogeneous mixing* within the population yields the relation

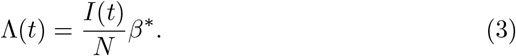

To simplify notation, we let *β* = *β*^*\**^*/N*. The formulation of the force of infection in (3) is referred to as *mass action transmission* [9]. The exposition above is extended to all susceptible individuals, leading to the continuous-time relation:

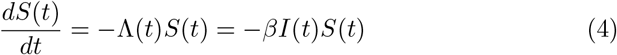

We start the discretization by fixing *h >* 0. By integrating (4) over the time interval (*t, t* + *h*], the following recurrence relation is deduced.

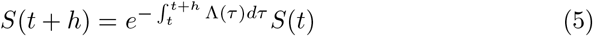

Equation (5) is now interpreted as the expected number of susceptible individuals at time *t*+*h*, when we assume that there are *S*(*t*) susceptible individuals at time *t*. We observe in turn that the first factor on the right hand side of (5) is the probability for a susceptible to escape from infection during the time interval (*t, t* + *h*]. Therefore, 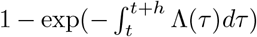 is the probability for a susceptible person to become infected during the time interval (*t, t* + *h*].

The following integral becomes the *cumulative force of infection* over (*t, t*+*h*].

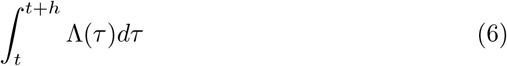

By taking the Taylor expansion of (6) around *t* and considering expressions (3), (5) together, the following holds.

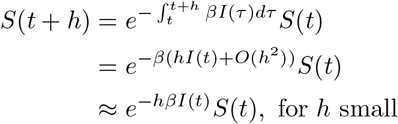

The probability *p*_1_(*t*) = 1 − *e*^−*hβI*(*t*)^ will now be regarded as the success probability for the Bernoulli trial corresponding to an interaction between an infectious and a susceptible individual, and the interaction occurs within (*t, t* + *h*]. By recalling the assumption of homogeneous mixing, the previous discussion suggests we define a random variable 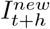 that follows a binomial distribution and which represents the number of newly infected individuals.

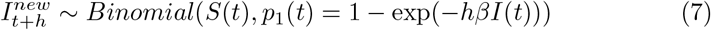

To finish this modelling part, the infectious period Δ(*t*) is assumed to be exponentially distributed with parameter *γ* ∈ R_≥0_, irrespective of the individual. Accordingly, the cumulative density function *p*_2_ = 1 – exp(–*hγ*) will represent the probability for an infectious individual to have recovered by time *h >* 0. This suggests that the corresponding number of newly recovered individuals 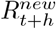 can be defined as a random variable with binomial distribution as follows.

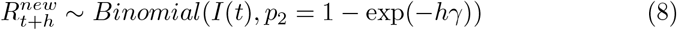

### 3.2. The SIR process

Let *S*(0), *I*(0), and *R*(0) be fixed constants such that *N* = *S*(0) +*I*(0) +*R*(0) — these are just the initial conditions for the ODE version of an SIR model — and *h >* 0. For all *t* ∈ ℕ, we define discrete random variables *S*_*t*_, *I*_*t*_, and *R*_*t*_, all of which take values from ℕ. In particular, let *S*_0_ = *S*(0), *I*_0_ = *I*(0), and *R*_0_ = *R*(0). For *t* ≥ 0 we base the following definition on Equations (7) and (8):

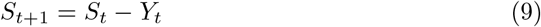

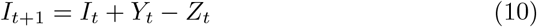

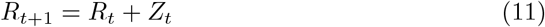

with

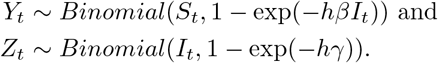

By definition, we have the property of *conservation of population*.

#### Lemma 2.

*For all t* ∈ ℕ *we have that N* = *S*_*t*_ + *I*_*t*_ + *R*_*t*_.

Let *t* ∈ ℕ be arbitrary and write (*S*_*t*_, *I*_*t*_, *R*_*t*_) = (*m*_1_, *m*_2_, *m*_3_). We will focus on the probability mass function 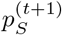 of *S*_*t*+1_. The equations below follow directly from our definition.

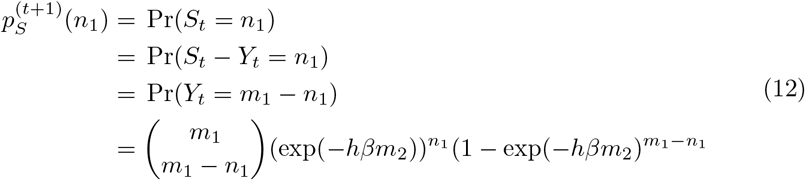

(Here, we adopt the convention that 0^0^ = 1 so that 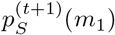 to be 1 when *m*_2_ = 0.) Similarly, for the probability mass function 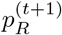 of R_t+1,_ we get the following equations.

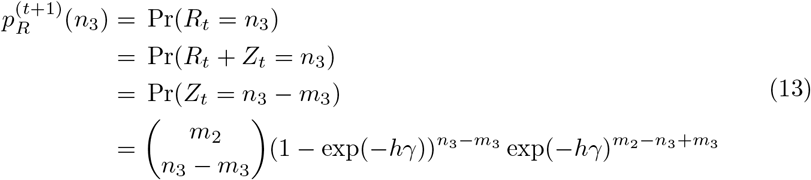

Observe, from (12) and (13), that *S*_*t*+1_ and *R*_*t*+1_ are independent. Recall that if *X* and *Y* are independent then Pr(*X* = *x, Y* = *y*) = Pr(*X* = *x*) Pr(*Y* = *y*) = *p*_*X*_ (*x*)*p*_*Y*_ (*y*), for *x, y* in the sample space. Thus, the joint probability mass function 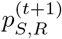 of (*S*_*t*+1_, *R*_*t*+1_) is given by 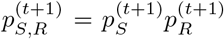. Finally, note that *I*_*t*+1_ = *N* − *S*_*t*+1_ − *R*_*t*+1_, by Lemma 2. Therefore, the joint probability mass function 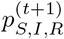 of (*S*_*t*+1_, *I*_*t*+1_, *R*_*t*+1_) is also given by 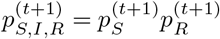.

Henceforth, we write *X*_*t*_ for (*S*_*t*_, *I*_*t*_, *R*_*t*_) and *Q* for the set of (*n*_1_, *n*_2_, *n*_3_) ∈ ℕ^3^ such that *N* = *n*_1_ + *n*_2_ + *n*_3_. For all *t* ∈ ℕ, *X*_*t*_ is a discrete random variable that takes values from *Q* and its probability mass function (as discussed above), for all *t* ≥ 1 is given by:

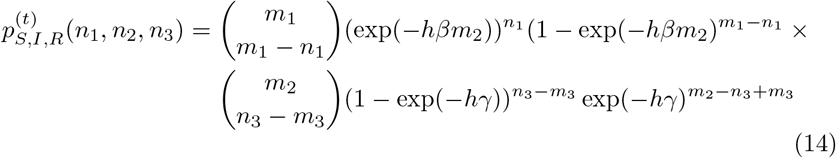

where *X*_*t*−1_ = (*m*_1_, *m*_2_, *m*_3_).

In the next section, we prove that (*X*_*t*_)_*t*≥0_ is indeed a stochastic process and even a Markov chain. For now, we can already define what will later become its transition patrix. First, note that the function from Equation (14) that the dependency on *t* can be changed to a dependency on ***m*** = (*m*_1_, *m*_2_, *m*_3_). Now, for all ***m, n*** = (*n*_1_, *n*_2_, *n*_3_) ∈ *Q*, we define:

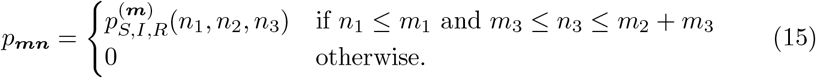

Then, the matrix ***P*** = (*p*_***mn***_ | ***m, n*** ∈ *Q*) is clearly stochastic. For convenience, we sometimes write ***P*** (***m, n***) instead of *p*_***mn***_.

To conclude this section we state the following property which follows immediately from the definition above.

#### Lemma 3.

*Let* ***m*** = (*m*_1_, *m*_2_, *m*_3_), ***n*** = (*n*_1_, *n*_2_, *n*_3_) ∈ *Q be states. Then*, ***P*** (***m, n***) *>* 0 *if and only if n*_1_ ≤ *m*_1_ *and m*_3_ ≤ *n*_3_ ≤ *m*_2_ + *m*_3_.

## 4. Properties of the SIR Stochastic Process

In this section we study the sequence (*X*_*t*_)_*t*≥0_ of random variables defined in the previous section. In particular, we show that it is indeed a stochastic process. Then, we argue that it is a Markov chain and that ***P*** is its transition matrix. To do this, we follow [12, Chapter 10] to explicitly construct the probability measure space it induces.

A *path* is a sequence *X*_0_ = ***m***_**0**_, *X*_1_ = ***m***_**1**_, … such that ***P*** (***m***_***j***_, ***m***_***j*+1**_) *>* 0 for all *j* ≥ 0. The set of all such sequences is referred to as *Paths*. In turn, *Paths*_*fin*_ denotes the set of path prefixes *X*_0_ = ***m***_**0**_, …, *X*_*n*_ = ***m***_***n***_, where *n* ≥ 0 and ***P*** (***m***_***j***_, ***m***_***j*+1**_) *>* 0 for 0≤*j < n*. The *length* of *x ∈ Paths*_*fin*_∈ (ℳ) is the function counting the number of elements in *x*, and we write ℓ(*x*) for short. Let *π* ∈ *Paths*, then 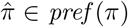 if 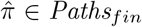 and 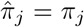 for 0 ≤ *j < n*, where 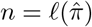. We introduce now the so-called *cylinder set* of 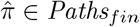:

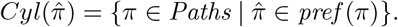

Consider a space (Ω, ℱ) with ℱ ⊆ *𝒫* (Ω). We say that a family *{E*_*j*_*}*_*j*≥1_ ⊆ ℱ *increases* to *E* and write *E*_*n*_ ↑ *E* if and only if:

- *E*_j_ ⊆*E*_*j+1*_, and
- ∪_*j* ≥1_ *E*_*j*_ = *E*.

We enunciate now Carathéodory’s extension theorem [13].

### Theorem 6.

*Let 𝒜 and σ*(*𝒜*), *be an algebra and its generated σ-algebra over a set* Ω. *If ν* : 𝒜 → ℝ_≥0_ ∪ *{*+∞*} is σ-additive, there is a σ-additive function π* : *σ*(𝒜) → ℝ_≥0_ ∪ *{*+∞*} such that π*|𝒜 = *v. Furthermore, if there exist {E*_*j*_*}*_*j*≥1_ ⊆ 𝒜 *such that E*_*j*_ ↑ Ω *and v*(*E*_*j*_) *<* ∞ *for all j* ≥ 1, *then π is unique*.

We use this last theorem to construct the probability measure space associated with (*X*_*t*_)_*t*≥0_. Let *n* ≥ 0 and let *A*_*n*_ ⊆ *Paths*_*fin*_ be as follows.

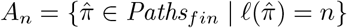

Take the family *{B*_*n*_*}*_*n*≥1_ given by 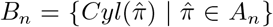. Let 𝒜 (∪_*n*≥1_ *B*_*n*_) be the algebra generated by ∪_*n*≥1_ *B*_*n*_.

### Lemma 4.

*Let ℱ be a semi-algebra over a set* Ω *and* 𝒜 (ℱ) *be the algebra generated by ℱ. If A* ∈ 𝒜 (ℱ), *there is a pairwise disjoint family* 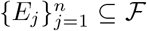 *such that* 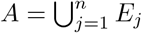.

Let *E* ∈ 𝒜 (∪_*n*≥1_ *B*_*n*_). There exist 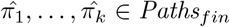, where the lengths of 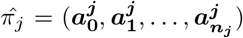) and 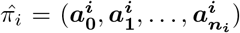 need not be the same for 1 ≤ *i, j* ≤ *k*, such that 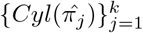 is a pairwise disjoint family and:

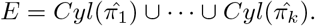

Let us define the function *μ* : *𝒜* (∪_*n*≥1_ *B*_*n*_) → ℝ_≥0_ ∪ *{*+∞*}* as follows.

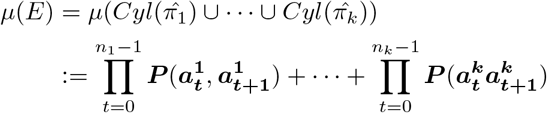

Note that this definition does cover all elements of 𝒜 (∪_*n*≥1_ *B*_*n*_). Indeed, consider a family *{E*_*j*_*}*_*j*≥1_ such that 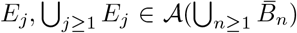 for all *j* ≥ 1 and *E*_*i*_ *∪ E*_*k*_ = ∅ when *i ?*= *k*. By Lemma 4, the sets *E*_*j*_ and *∪* _*j*≥1_ *E*_*j*_ can always be expressed as a finite union of pairwise disjoint cylinder sets. Taking this into account allows us to evaluate *μ* of ∪_*j* ≥1_ *E*_*j*_ and *E*_*j*_ as (the limit of) a sum, as defined above.

Let *σ*(∪_*n*≥1_ *B*_*n*_) denote the *σ*-algebra generated by ∪_*n*≥1_ *B*_*n*_. uWe use The-orem 6 to show the existence of a *σ*-additive function *θ* : *σ*(∪_*n*≥1_ *B*_*n*_) → ℝ_≥0_ ∪ *{*+∞*}* such that 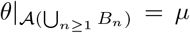. We prove now that *θ* is a probability function and that *μ* is *σ*-finite. Let ***m*** = (*S*(0), *I*(0), *R*(0)) be the initial conditions for the ODE from the previous section — that is, *X*_0_ = ***m*** in the sequence of random variables we have defined. Note that Ω = *Paths* follows the next decomposition.

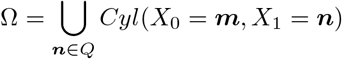

Therefore *θ*(Ω) = *μ*(Ω) = Σ_***n***∈*Q*_ ***P*** (***m, n***). Since ***P*** is a stochastic matrix, it follows that *θ*(Ω) = 1. Now, define *E*_*j*_ ∈ 𝒜 (∪_*n*≥1_ *B*_*n*_) and *Q*_*j*_ ⊆ *Q* for 1 ≤ *j* ≤ |*Q*| inductively as follows. We set *Q*_1_ = *Q\{****n****}* and *E*_1_ = *Cyl*(*X*_0_ = ***m***, *X*_1_ = ***n***) for an arbitrary ***n*** ∈ *Q*. For *j >* 2, let ***n*** ∈ *Q*_*j*−1_ and set *Q*_*j*_ = *Q*_*j*−1_ *\ {****n****}, E*_*j*_ = *E*_*j*−1_ ∪ *Cyl*(*X*_0_ = ***m***, *X*_1_ = ***n***). Note that *E*_*j*_ ↑ Ω and *μ*(*E*_*j*_) *<* ∞, for *j* ≥ 1. Thus, Ω is *σ*-finite and the extension *θ* of *μ* is unique.

### Theorem 7.

*The SIR process* (*X*_*t*_)_*t*≥0_ *is a stochastic process. Moreover, it is a Markov chain with state space Q transition matrix* ***P***.

Proof. By the above arguments, the SIR process (*X*_*t*_)_*t*≥0_ is a stochastic process with probability space (*Paths, σ*(∪ _*n*≥1_ *B*_*n*_), *θ*). Since *μ* satisfies the Markov property by definition, *θ* does so too by extension. It then follows from Theorem 6 that the SIR stochastic process is also a Markov chain. To conclude, by definition of *μ*, we get that the transition matrix of the Markov chain is ***P***.

To keep the notation simpler and uniform, in the rest of this work we write (Ω,, Pr) for the probability space (*Paths, σ*(∪ _n≥1_ *B*_*n*_), *θ*) defined in this section.

### 4.1. The Markovian SIR model

In this section we study the SIR Markov chain (*X*_*t*_)_*t*≥0_ defined in the previous sections. We take the initial distribution ***λ*** given by *δ*_***m***_, where ***m*** = (*N* −1, 1, 0), i.e., with probability one, (*X*_*t*_)_*t*≥0_ starts off from the state corresponding to one infectious individual in a population of otherwise susceptible individuals.

### 4.2. The SIR Markov chain is absorbing

We start by arguing that the SIR Markov chain is absorbing. This will enable the results we have recalled in Section 2 for absorbing Markov chains. First note that by Lemma 8 the set:

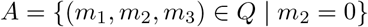

is nonempty and that it is exactly the set of absorbing states of the SIR Markov chain. It remains for us to prove that *A* is reachable from all states.

Observe that for all ***m*** = (*m*_1_, *m*_2_, *m*_3_) ∈ *Q* there exists ***n*** = (*n*_1_, 0, *n*_3_) ∈ *A* such that *n*_1_ ≤ *m*_1_ and *m*_3_ ≤ *n*_3_. Let ***m***^***′***^ = (*n*_1_, *m*_2_ + *m*_1_ − *n*_1_, *m*_3_) and note that ***m***^***′***^ ∈ *Q*. By Lemma 3, we have that ***P*** (***m, m***^***′***^), ***P*** (***m***^***′***^, ***n***) *>* 0 hence:

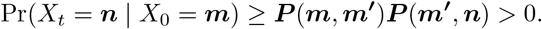

#### Lemma 5.

*The SIR Markov chain* (*X*_*t*_)_*t*≥0_ *is absorbing*.

### 4.3. The Expected Epidemic Duration

In this section we address the first hitting time to the set of absorbing states *A* of the SIR Markov chain. We denote the expected hitting time to this specific set by E[*τ*_end_], which in turn can be computed by using the expression in Theorem 3. Instead, we take a first step towards optimizing this computation by considering Lemma 3, which gives a characterization of the nonzero terms in ***P***. In this way, for the specific case of the SIR Markov chain, the calculations from Theorem 3 are reduced to what is shown in the next lemma.

#### Lemma 6.

*For all states* ***m*** ∈ *Q, we have that* 𝔼_***m***_[*τ*_end_] *is:*

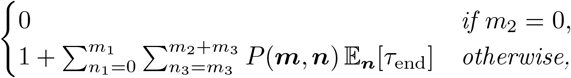

*where* ***n*** = (*n*_1_, *N* − *n*_1_ − *n*_3_, *n*_3_).

In the following section we turn our attention to concrete algorithms that allow us to compute the nonzero expressions from Lemma 6. We remark that the non-linearity in the original deterministic account of the contact process (see Equation (4)) is not to be seen in the formulation of our research problem (given by Lemma 6). This will simplify the analysis, as it will be seen in the next section.

## 5. Algorithms for the Expected Epidemic Duration

As a first algorithm, we will specialize Theorem 5 to our SIR Markov chain leveraging Lemma 6. Later, we give pseudo-code for the algorithm described and develop a non-trivial optimization to make it faster (by a linear factor).

### 5.1. Linear algebra-based approach

We will rewrite the expression corresponding to the non-absorbing states in Lemma 6 using matrix algebra. In doing so, we obtain a mathematical form that already illustrates the guiding principle behind the algorithmic design we use to compute the vector of mean hitting times later. For this purpose, the states of the system are ordered according to the relation:

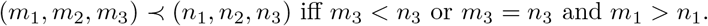

Let the transient states *Q \ A* of the Markov chain be numbered from one to *t* according to the ≺-relation: (***m***_**1**_, ***m***_**2**_, …, ***m***_***t***_). Write ***k***^***A***^ for the vector of mean hitting times 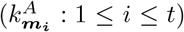 corresponding to those *t* states in that same order. By Theorem 5, we have ***k***^***A***^ = ***N* 1**. By definition of ***N*** we thus get the following relation.

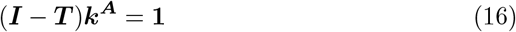

Now, using the order we imposed on the transient states together with Lemma 3, we can show the following.

#### Lemma 7.

*Consider the SIR Markov chain* (*X*_*t*_)_*t*≥0_. *The matrix* ***I*** − ***T*** *is upper triangular*.

This enables the computation of ***k***^***A***^ by means of *backward substitution*.

In general, for upper triangular systems, i.e., systems of the form ***Ux*** = ***b***, where ***x, b*** ∈ ℝ^*m*^ and ***U*** is an invertible upper triangular matrix of size *m* × *m*, the back substitution algorithm is given in [14] and it requires *m*^2^ elementary operations: substitution and solving for a variable. Now, the size of the system (16) is (*N* + 1)(*N* + 2)*/*2, and therefore the application of back substitution to our system has a worst-case time complexity of *O* (*N* ^4^), provided ***P*** has been pre-computed. In the next section, we instantiate this approach in the form of a concrete algorithm. (We thus also explicitly compute ***P*** and analyze the complexity of doing so.) Then, we give a nontrivial optimization based on dynamic programming to compute the transition matrix ***P*** efficiently.

#### Algorithm 1

A simple algorithm to compute 𝔼^***m***^[*τ*_end_] from all ***m***

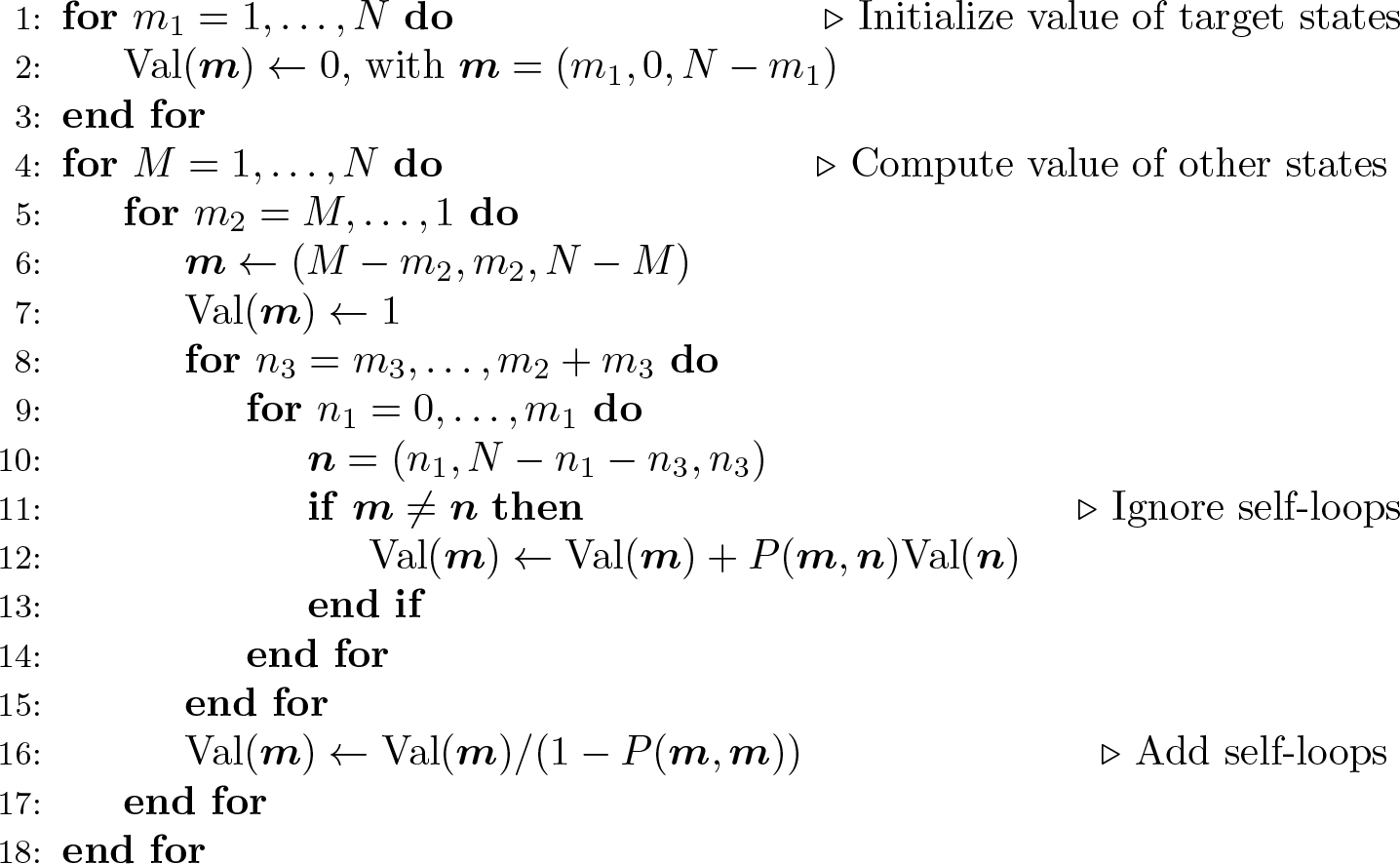

### 5.2 Dynamic programming solution

Algorithm 1 exploits equalities from Lemma 6 in order to compute the expected hitting times from all states. Notice that, by conservation of population, two for-loops suffice to go through all source and target states. That is, in line 10 no further for-loop is needed since there is a unique value of *n*_2_ that makes (*n*_1_, *n*_2_, *n*_3_) a valid state. Further note that in line 4 we cycle through all values of *m*_1_ + *m*_2_ which is why the following for-loop runs until *M*. Then, similar to the case of ***n***, no further for-loop is needed since there is a unique value of *m*_3_ such that (*M* − *m*_2_, *m*_2_, *m*_3_) is a valid state.

The next property, which follows directly from Lemma 3, will be useful in our analysis of Algorithm 1.

#### Lemma 8.

*Let* ***m, n*** *be states. If P* (***m, n***) *>* 0 *then:*

- *either* ***m*** = ***n*** *or*
- *m*_1_ ≥ *n*_1_, *m*_1_ + *m*_2_ ≥ *n*_1_ + *n*_2_, *and one of the latter two is strict*.

Note that Algorithm 1 clearly terminates. Further observe that the algorithm has 4 nested for-loops, each ranging over at most *N* values each. Since *P* (***m, n***) can be calculated using *O* (*N*) elementary operations^1^ using Equation(14), we get the following more precise statement.

#### Theorem 8 (Complexity)

*The worst-case time complexity of Algorithm 1 is O* (*N* ^5^).

Regarding the values that the algorithm computes, we can prove that they are indeed the required expected hitting times.

Regarding the values that the algorithm computes, we can prove that they are indeed the required expected hitting times.

#### Theorem 9 (Correctness)

*Let* Val(·) *be as computed by Algorithm 1. Then*, Val(***m***) = 𝔼^***m***^[*τ*_end_] *for all states* ***m***.

Proof. First, note that Lemma 8 guarantees Val(***n***) has already been computed when used in line 12. Second, we can focus on Val(***m***) such that *m*_2_ *>* 0 since for the remaining ***m***, the claim holds already after the initialization ends in line 3.

Now, for an arbitrary ***m*** as computed in line 6 let Val_1_(***m***) denote the value of Val(***m***) after line 7 is executed; Val_2_(***m***) its value after line 15 is executed; and Val_3_(***m***) its value after line 16. We will argue that Val_3_(***m***) = 𝔼^***m***^[*τ*_end_], which will conclude the argument because Val(***m***) is not updated afterwards. Note that the following hold.

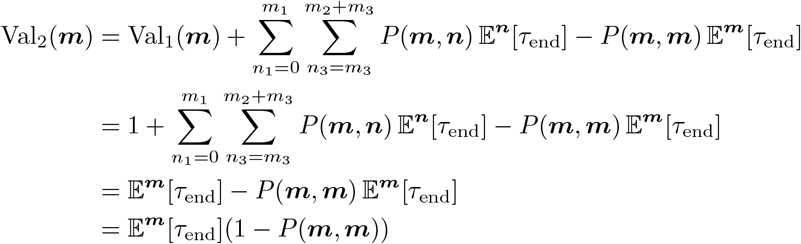

Finally, since Val_3_(***m***) = Val_2_(***m***)*/*(1*− P* (***m, m***)), we get that Val_3_(***m***) = 𝔼^***m***^[*τ*_end_].

The question is now: can one do better? In the next section we give a positive answer to this question by improving on our algorithm using dynamic programming to efficiently pre-compute the values *P* (***m, n***) for all ***m*** and ***n***.

### 5.3. A more efficient algorithm

Recall Equation (14) gives us that for all states ***m*** ≠ ***n*** such that *n*_1_ ≤ *m*_1_ and *m*_3_ ≤ *n*_3_ ≤ *m*_2_ + *m*_3_ the probability *P* (***m, n***) of transitioning from ***m*** to ***n*** is as follows.

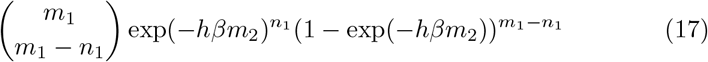

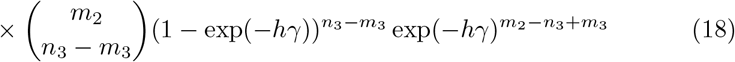

The crux of our improvement on Algorithm 1 is the following recurrence.

#### Lemma 9.

*Let* ***m, n*** *be states such that n*_1_ *< m*_1_ *and m*_3_ *< n*_3_ ≤ *m*_2_ + *m*_3_. *Then, P* (***m, n***) = *α*(***m, n***)*P* (*m*_1_ *−* 1, *m*_2_, *m*_3_ + 1, ***n***), *where:*

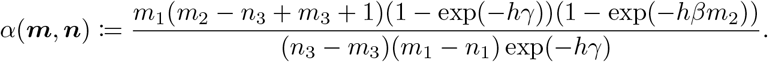

Proof. Note that *P* (***m, n***), *P* (*m*_1_ *−* 1, *m*_2_, *m*_3_ + 1, ***n***) *>* 0 by Lemma 3 and our assumptions. We first consider the binomial coefficients of (17) and (18).

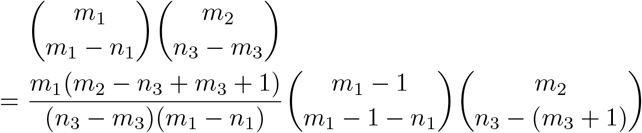

#### Algorithm 2

An efficient algorithm to compute *P* (***m, n***) for all ***m*** and ***n***

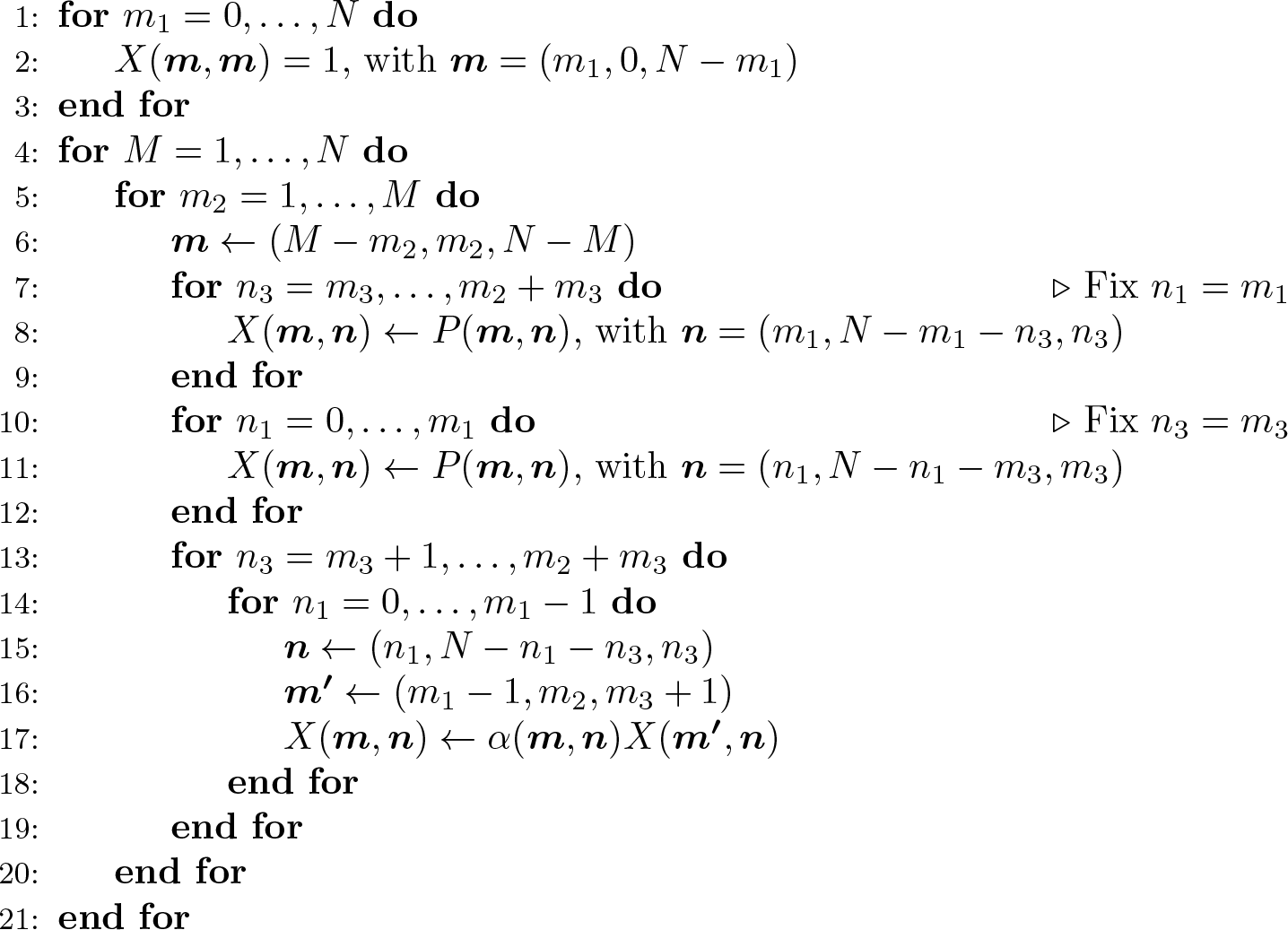

Importantly, since we have assumed that *m*_3_ *< n*_3_ and *n*_1_ *< m*_1_, the denominator of the fraction above is not 0. On the other hand, for the terms involving “probabilities” — that is, exponential terms — we observe that the following is true for (17).

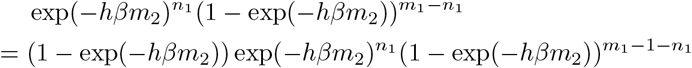

Similarly, for the probabilities in (18) we note the following.

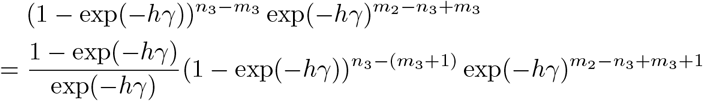

Indeed, *α*(***m, n***) is exactly the product of the coefficients on the right-hand sides of the above equalities. This concludes the proof as the product of the left-hand sides is exactly *P* (***m, n***) and that of the right-hand sides (after factoring out *α*) is *P* (*m*_1_ *−* 1, *m*_2_, *m*_3_ + 1, ***n***). ?

Based on Lemma 9, we propose Algorithm 2 to compute *P* (***m, n***) for all ***m*** and ***n***. The recurrence relation allows us to compute new transition probabilities based on previously computed ones in a manner reminiscent of how Pascal’s rule is used to construct Pascal’s triangle.

As with Algorithm 1, we observe that Algorithm 2 clearly terminates because all for-loops are bounded and there are no jump statements in the code. Regarding the time complexity of Algorithm 2, however, we now only ever nest at most 4 for-loops and *P* (***m, n***) is never used (i.e. computed) within a for-loop nesting of depth 4.

#### Theorem 10 (Complexity)

*The worst-case time complexity of Algorithm 2 is O* (*N* ^4^).

For correctness, we have the following statement.

#### Theorem 11 (Correctess)

*Let X* (·) *be as computed by Algorithm 2. Then, X* (***m, n***) = *P* (***m, n***) *for all pairs of states* ***m, n*** *such that n*_1_ ≤ *m*_1_ *and m*_3_ ≤ *n*_3_ ≤ *m*_2_ + *m*_3_.

Proof. First, one needs to take care that the previous transition probabilities must be defined before they are being used. Note that this is only relevant in lines 13–19. It thus follows from the order in which the states ***m*** are traversed that *X* (***m***^***′***^, ***n***) has already been computed in a previous iteration.

Now, it is easy to see that *X*(***m, n***) = *P* (***m, n***) if either *m*_1_ = *n*_1_ or *m*_3_ = *n*_3_ since lines 8 and 11 literally assign the right-hand side to the left-hand side of the equality. If neither equality holds, then *X*(***m, n***) is computed in the loop 13–19 and *X*(***m, n***) = *P* (***m, n***) by Lemma 9. Indeed, the conditions that *n*_1_ *< m*_1_ and *m*_3_ *< n*_3_ are guaranteed by values over which *n*_3_ and *n*_1_ range in the loops from lines 13 and 14.

Together with the theorems regarding Algorithm 1, the above imply the following.

#### Theorem 12.

*Algorithm 1, when pre-computing P* (***m, n***) *using Algorithm 2, computes* 𝔼^***m***^[*τ*_end_] *for all states* ***m*** *in time O* (*N* ^4^).

## 6. Experiments and Conclusions

In this section we test Algorithm 1 and Algorithm 2 against the use of simulations to estimate mean epidemic durations. All the code used to generate the experiments we present here can be fetched from https://github.com/jalarcon-ale/MarkovChain.

In Figure 3 we have plotted sample means of simulation experiments. Each experiment corresponds to the duration of a simulation of the SIR Markov chain, taking as initial condition the state corresponding to one infectious individual in an otherwise susceptible population. The population size has been fixed to *N* = 50, and the number of experiments per sample increases by ten, starting from one, and ending with 10001. The plot suggests that using six thousand simulated durations of the SIR Markov chain yields a good estimate of the mean epidemic duration. We remark that a correct simulation study aimed at estimating the mean duration should follow a methodology (see for instance [15] and [16]). One of the advantages of using the method developed in this paper is that such analysis can be avoided.

**Figure 2:**
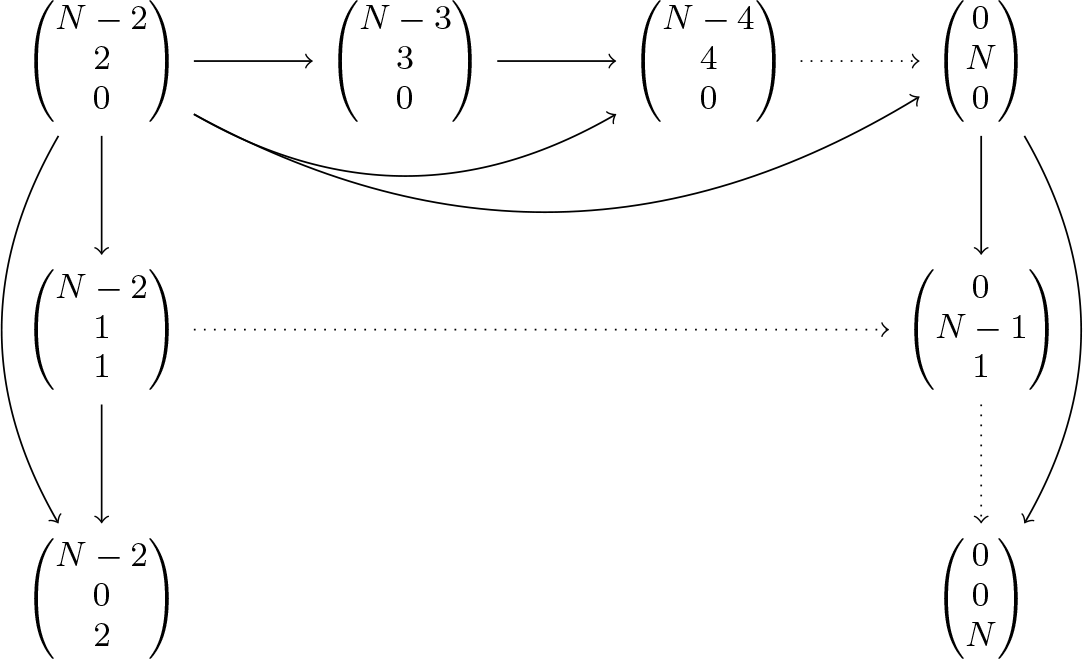
The SIR Markov chain has the interesting property that each state has a very large number of successor states. So much so that from any state one can reach an absorbing state in at most two transitions.

**Figure 3:**
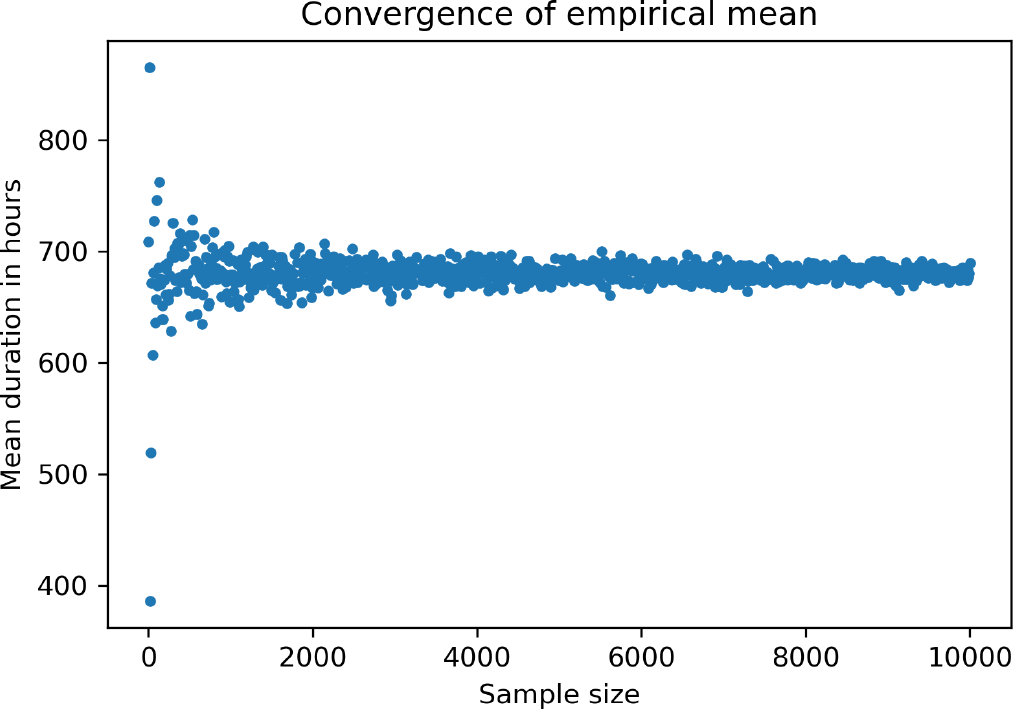
Converging behavior of the sample mean when the number of experiments increases. The sample size indicates the number of simulations of the SIR Markov chain used to estimate the mean epidemic duration. The population size has been fixed to *N* = 50, and the parameter values used are *β* = 1*/*(2*N*) and *γ* = 1*/*6, while the time step *h* is one hour.

To empirically validate Algorithm 1 and Algorithm 2 we have performed 100 thousand simulations of the SIR Markov chain, with fixed parameters and for ten population sizes, ranging from ten to 100 individuals. In Figure 4a the corresponding durations are represented with *boxplots*^2^. In Figure 4b, the empirical means are illustrated with red dots, and Algorithm 1 and Algorithm 2 were used together to compute the corresponding exact mean epidemic durations, represented by blue dots.

**Figure 4:**
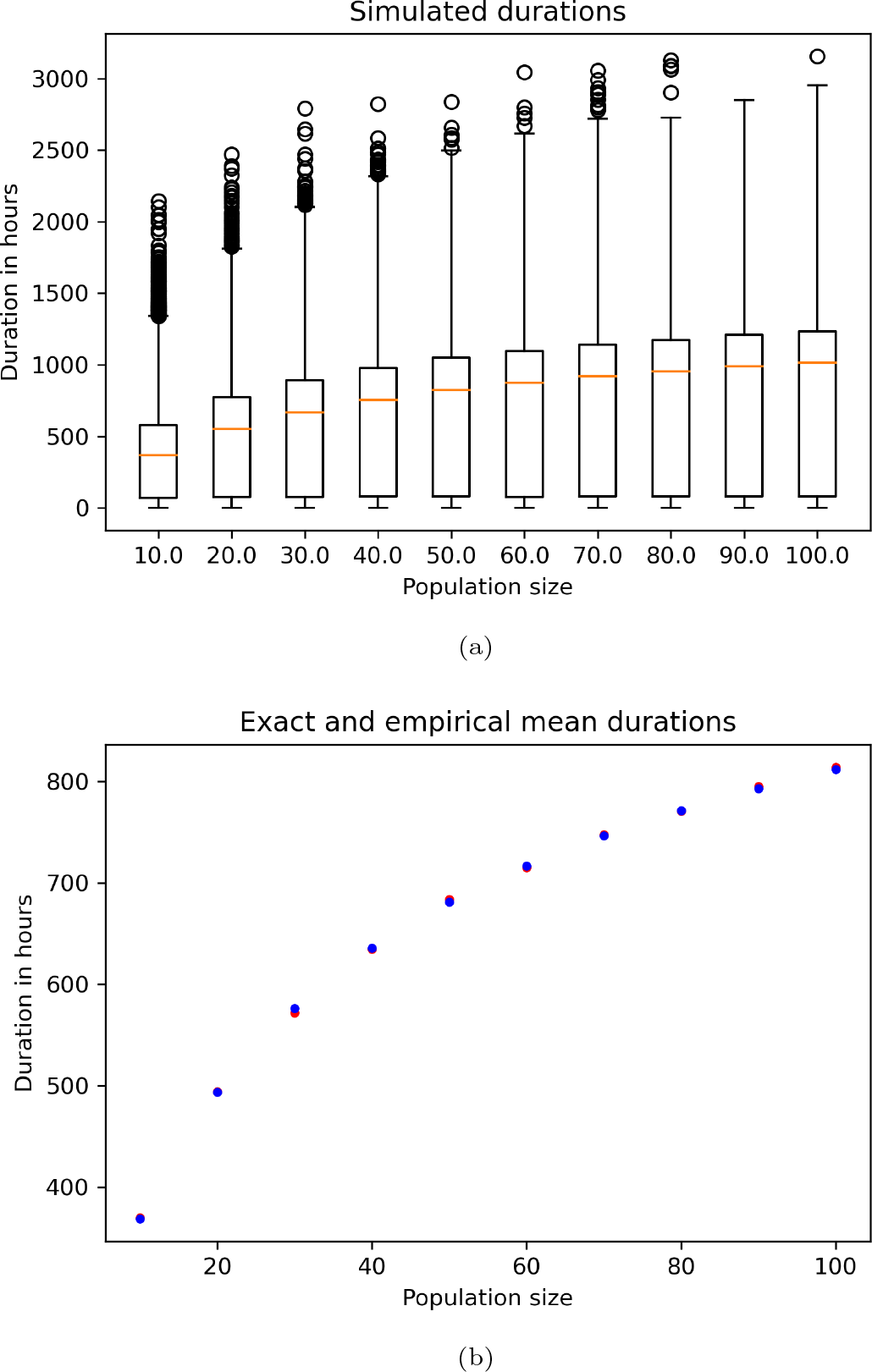
For a range of population sizes *N* varying from ten to 100 individuals, Figure 4a shows 100 thousand simulated durations of the SIR Markov chain. In Figure 4b the corresponding empirical means (red dots) are plotted together with their exact counterparts (blue dots). The parameter values used are *β* = 1*/*(2*N*) and *γ* = 1*/*6, while the time step *h* is one hour.

### 6.1. Future work

The methodology from this work can be extended to models with more compartments and with contact structure. However, for models with branching, i.e., models where an individual has more than one possibility to change compartments, we remark that the derivation of the transition matrix poses its own challenge. Namely, branching yields case definition and every sub-branch will add a corresponding sub-case. The chain of conditionals involved in this process carries a rather cumbersome bookkeeping effort with itself. For this kind of models, (algorithmic) automation of the transition-matrix derivation should be addressed as a first step towards the extension of our method.

## Data Availability

All data produced are available online at https://github.com/jalarcon-ale/MarkovChain

## Acknowledgments

We would like to thank Niel Hens for very useful feedback on early version of this work, Kasper Engelen and Shrisha Rao for proof-reading the article.

Financial support for this work was provided by the Flemish inter-university (iBOF) “DESCARTES” project.

## Footnotes

1 We take binary addition and multiplication to be elementary operations. Hence, while exponentiation can be realized using a logarithmic number of such operations via iterated squaring, we are not aware of an algorithm to compute factorials using a sub-linear number of multiplications.

2 These are plots that summarize statistical properties of a data set. Concretely, a boxplot creates a box from the first quantile to the third quantile, and a (red) segment of line at the median.

## Notes

### Competing Interest Statement

The authors have declared no competing interest.

### Summary of Updates

Figures 3 and 4 revised; Section 6 updated to account for Figures 3 and 4; Author list updated.

